# A liver-heart endocrine axis revealed by systems genetics and mediated by hepatocyte growth factor activator

**DOI:** 10.64898/2026.05.05.26352474

**Authors:** Michal Juda, Dylan Sarver, Jenny Cheng, James R. Hilser, Xinmin S. Li, Tomohiro Yokota, Christopher Li, Calvin Pan, Zhiqiang Zhou, Adrian Arrieta, Marcus Seldin, Xia Yang, W H Wilson Tang, Thomas M. Vondriska, Stanley L. Hazen, Hooman Allayee, Aldons J. Lusis

**Author notes:** **Correspondence**: Aldons J. Lusis. **Conflict-of-interest**: Dr. Hazen reports being named as co-inventors on pending and issued patents held by the Cleveland Clinic relating to cardiovascular diagnostics and therapeutics and being eligible to receive royalty payments for inventions or discoveries related to cardiovascular diagnostics or therapeutics from Cleveland HeartLab, Quest Diagnostics and Procter & Gamble. Dr. Hazen also reports being a paid consultant for Zehna Therapeutics and having received research funds from Zehna Therapeutics. The other authors declare no competing interests.

## Abstract

The liver and heart are tightly interconnected organs, and liver disease is frequently accompanied by cardiovascular dysfunction, including heart failure^1–5^. Despite this clinical association, the mechanisms by which liver-derived endocrine signals influence cardiac gene programs and disease susceptibility remain poorly defined. Inter-organ endocrine communication is increasingly recognized as a key regulator of systemic physiology, including cardiac function^6–8^, but a comprehensive understanding of liver–heart communication is lacking. Here we use an unbiased, population-based systems genetics approach in a genetically diverse mouse cohort to identify liver-derived secreted factors associated with cardiac transcriptomic variation. This analysis reveals hepatocyte growth factor activator (HGFAC) as a candidate mediator of inter-organ communication. Cross-tissue analysis of human genetic and transcriptomic datasets further suggests a conserved relationship between hepatic HGFAC expression and cardiac gene programs. These observations implicate a previously unrecognized liver–heart axis that appears to contribute to heart failure pathophysiology across species.

## Main

Liver–heart communication is increasingly recognized as an important component of systemic cardiovascular physiology. The liver secretes a wide range of endocrine and paracrine factors that regulate metabolism^9^, inflammation^10^, and vascular function^11^; however, how liver-derived signals coordinate cardiac gene expression programs across physiological and disease contexts remains incompletely understood. It is not known whether specific hepatically secreted proteins are linked to coordinated transcriptional programs in the heart in a conserved manner across species.

To address this, we sought to systematically identify liver-derived secreted factors associated with cardiac transcriptomic variation using an unbiased systems genetics framework. We leveraged the genetically diverse Hybrid Mouse Diversity Panel (HMDP), which enables mapping of naturally occurring variation in hepatic gene expression to global changes in cardiac gene expression across strains^12,13^. This approach allows identification of candidate endocrine mediators based on cross-organ transcriptional coupling rather than predefined pathway selection. Among liver-enriched secreted genes, HGFAC^14^, a hepatocyte-specific serine protease that converts pro–hepatocyte growth factor (pro-HGF) into active HGF^15,16^, emerged as a top candidate associated with cardiac gene expression modules (**Fig. 1a, Extended Data Fig. 1**). This association suggested a potential link between hepatic protease activity and coordinated cardiac transcriptional programs, consistent with clinical evidence that circulating HGF predicts mortality in patients with heart failure^17–19^. HGFAC, HGF, and MET, the HGF receptor, are all highly conserved across mouse, rat, and human (**Extended Data Fig. 2**), consistent with evolutionary conservation of the HGFAC–HGF–MET signaling axis.

**Fig. 1.**
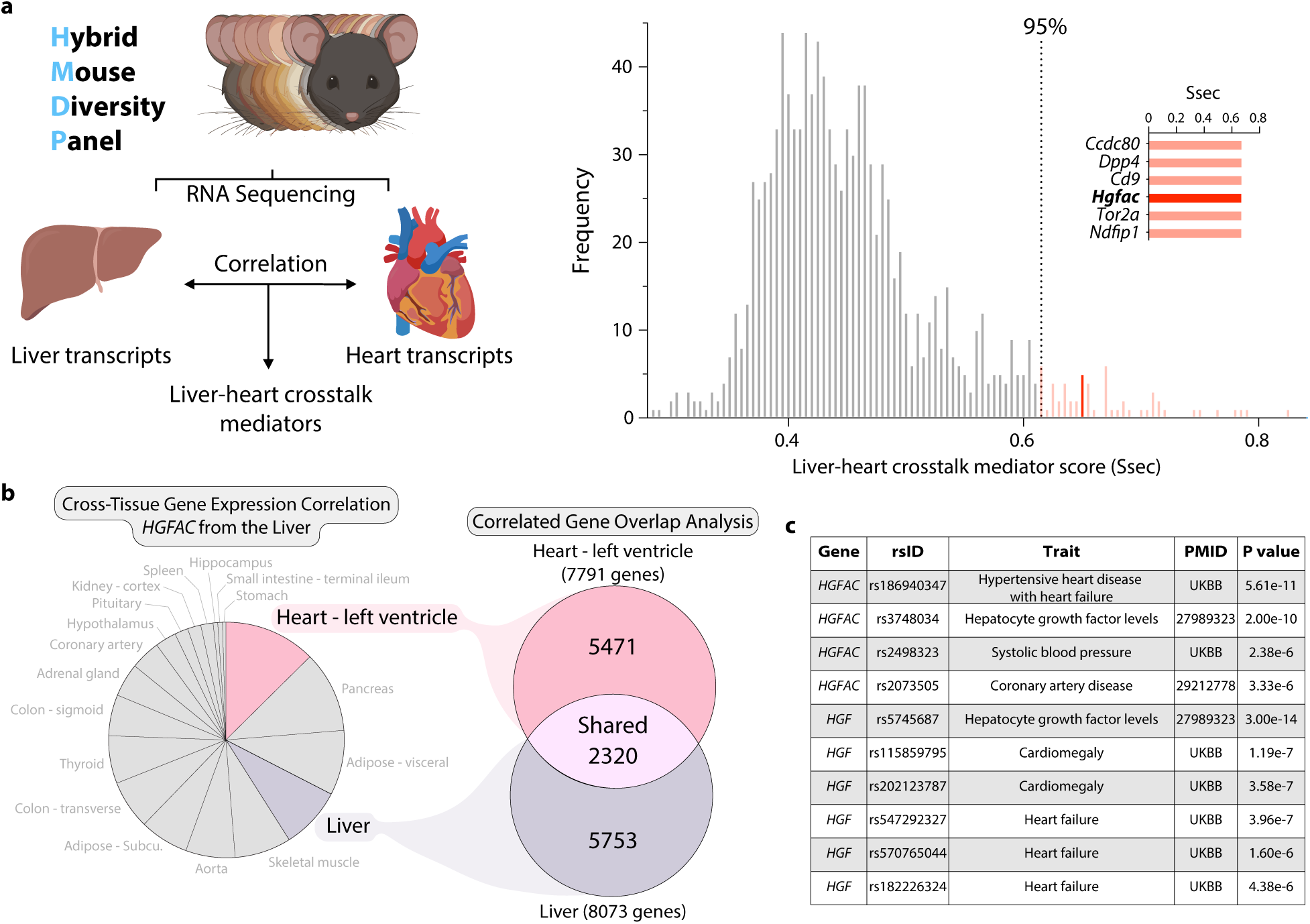
Identification of a hepatic HGFAC–driven liver–heart signaling circuit in mice and humans. **(a)** Schematic illustrating the analytical framework used to identify liver–heart crosstalk mediators by integrating bulk RNA sequencing data and cross-tissue transcript correlation analysis. Liver–heart crosstalk scores (S_sec_) calculated using QENIE, which quantifies the aggregate correlation (biweight midcorrelation) between the expression of a liver-expressed secreted protein gene and the global cardiac transcriptome, identify *Hgfac* as a candidate liver-derived mediator. (**b**) Expression correlation analysis using GD-CAT to assess data from the Genotype-Tissue Expression (GTEx) project where tissues were assessed with the expression of *HGFAC* in liver tissue with a qvalue cutoff of q<0.1. All 310 individuals (across both sexes) were used, and FDR adjustments were calculated using a Benjamini-Hochberg method. (**c**) List of disease and traits associated with *HGFAC* and *HGF* genetic variants in humans using PhenoScanner V2.

To determine whether this liver-heart transcriptional crosstalk is also conserved in humans, we applied gene-derived cross-tissue correlation analysis (GD-CAT^20^) to GTEx^21^ data. Hepatic HGFAC expression showed its strongest correlation with left ventricular gene expression programs, indicating a conserved inter-organ transcriptional relationship rather than a liver-restricted coexpression signature (**Fig. 1b**). To further assess relevance in human cardiovascular disease, we examined genetic association data across large cohorts. Variants near *HGFAC* and *HGF* were associated with cardiac remodeling phenotypes, including hypertensive heart disease, coronary artery disease, cardiomegaly, and heart failure (**Fig. 1c**). Distinct variants at both loci were also associated with circulating HGF levels, suggesting that genetic variation at *HGFAC* and *HGF* can modulate HGF abundance. Together, these analyses identify HGFAC as a liver-derived factor linked to coordinated cardiac gene expression programs across mouse and human systems, suggesting the presence of a previously unrecognized liver–heart endocrine axis.

We next asked whether activation of the HGFAC–HGF axis is associated with heart failure pathophysiology *in vivo*. To test this in mice, we examined a “two-hit” model of HFpEF combining high-fat diet and Nω-nitro-L-arginine methyl ester (L-NAME), which induces metabolic and vascular stress while preserving systolic function^22^. Circulating levels of both HGFAC and HGF were elevated in HFpEF mice compared to controls (**Extended Data Fig. 3**), suggesting activation of this signaling axis in disease states relevant to HFpEF. To determine whether increased hepatic HGFAC is sufficient to modify cardiac function *in vivo*, we selectively overexpressed *Hgfac* in hepatocytes using AAV8-mediated gene delivery under a hepatocyte-specific promoter. This approach produced a sustained increase in circulating HGFAC levels (**Extended Data Fig. 4**). Mice with hepatic HGFAC overexpression exhibited exacerbated HFpEF phenotypes, including increased diastolic dysfunction, reduced exercise capacity, and cardiac hypertrophy, while left ventricular ejection fraction remained preserved (**Extended Data Fig. 5**). Importantly, hepatic HGFAC overexpression in otherwise healthy mice did not produce detectable changes in cardiac structure or function (**Extended Data Fig. 6**), indicating that its effects are context-dependent and emerge in the presence of metabolic or vascular stress. Together, these findings indicate that hepatic activation of the HGFAC axis exacerbates HFpEF phenotypes *in vivo*, identifying this liver-derived pathway as a context-dependent driver of disease susceptibility rather than baseline cardiac function.

To define the cardiac mechanisms underlying HGFAC-associated HFpEF phenotypes, we performed transcriptomic and cell-type–resolved analyses of hearts from mice subjected to the HFpEF diet and hepatic *Hgfac* overexpression. Bulk RNA sequencing identified broad transcriptional changes enriched for receptor tyrosine kinase signaling pathways, consistent with activation of HGF–c-MET signaling downstream of HGFAC (**Extended Data Fig. 7**). We next performed single-nucleus RNA sequencing in HFpEF mice with hepatic *Hgfac* overexpression to identify the primary responding cardiac cell populations. Cardiomyocytes exhibited the most pronounced transcriptional remodeling among all cardiac cell types, indicating that they are the principal effector population of this axis (**Fig. 2a-c**). Cell–cell communication analysis (CellChat^23^) further revealed selective strengthening of cardiomyocyte–endocardial and cardiomyocyte–macrophage interactions following *Hgfac* overexpression (**Fig. 2d-f**). In contrast, global intercellular communication within the ventricle was reduced, with decreases in both the number and strength of predicted interactions across multiple cell types (**Fig. 2g-h**). Together, these findings indicate that hepatic HGFAC signaling is associated with a reorganization of cardiac intercellular communication networks in HFpEF, characterized by cardiomyocyte-centric signaling reinforcement alongside widespread attenuation of ventricular cellular connectivity.

**Fig. 2.**
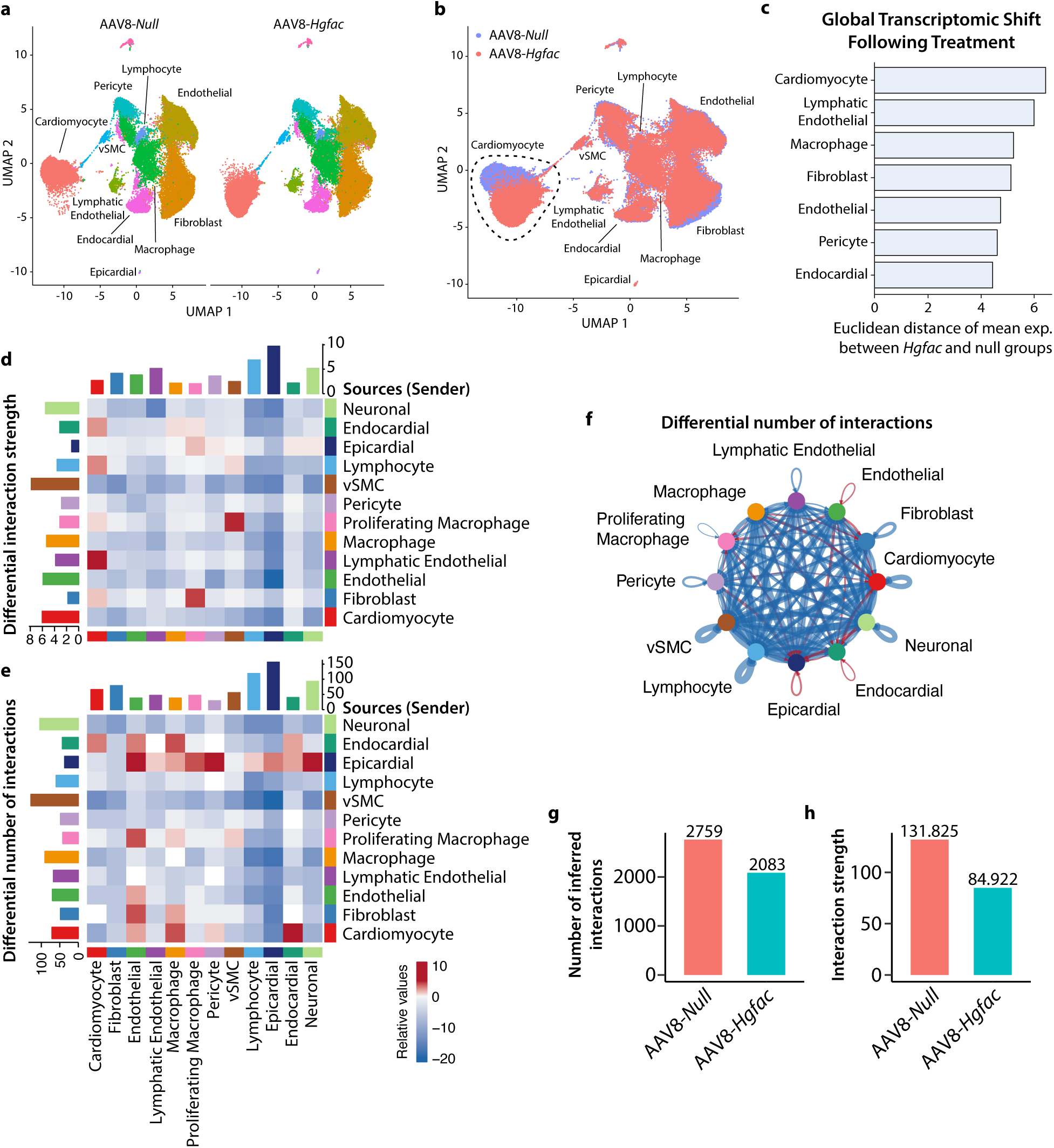
*Hgfac* overexpression alters intercellular communication in the HFpEF heart. 8-week-old C57BL/6J male mice were injected with of AAV8-TBG-*Hgfac* or AAV8-*Null* and were supplemented HFD and L-NAME for 7 weeks. (**a and b**) UMAP plot of single-nuclei profiles (dots) from 10X libraries colored by (**a**) cell-type and (**b**) treatment condition. The cardiomyocyte population is outlined in black. (**c**) Euclidean distance between average gene expression profiles of *Null* and *Hgfac* groups from (**b**) was calculated to quantify treatment-associated transcriptional divergence. (**d-h**) CellChat objects were generated using all cardiac cell types identified from HFpEF snRNA-seq (*Null* vs. *Hgfac*). (**d and e**) The differential interaction strength and number of interactions of inferred cell-cell communications are presented as a heat map listing the source cell (sender) on the Y-axis. (**f**) Differential number of interactions depicted as a circle plot. (**g**) Number of inferred interactions and (**h**) the interaction strength based on condition (*Null* vs *Hgfac*).

To determine whether HGFAC-axis signaling directly interacts with cardiomyocytes, we next examined neonatal rat ventricular myocytes (NRVMs) as a reductionist *in vitro* model using hypertrophy as a functional readout. Because HGFAC functions upstream of HGF, we tested whether downstream ligands of this axis directly modulate cardiomyocyte growth responses. NRVMs were stimulated with phenylephrine to induce hypertrophic stress and treated with recombinant HGF, HGFAC, or macrophage-stimulating protein (MSP^24^), an HGF-like ligand (**Fig 3a**). Among these factors, only HGF potentiated phenylephrine-induced cardiomyocyte hypertrophy in a dose-dependent manner, whereas HGFAC and MSP had no effect relative to NRVMS that were not treated with any recombinant protein (**Fig 3b**). To determine whether this effect was mediated through canonical receptor signaling, NRVMs were co-treated with HGF and the clinically approved c-MET inhibitor tepotinib^25^. Pharmacologic inhibition of c-MET fully abrogated HGF-induced hypertrophy, indicating that HGF acts directly on cardiomyocytes through c-MET-dependent signaling to promote cellular growth responses (**Fig 3c-d**). We next evaluated whether pharmacologic inhibition of c-MET could modify HFpEF development *in vivo*. In mice subjected to the two-hit HFpEF model, treatment with tepotinib attenuated cardiac hypertrophy, improved diastolic dysfunction, and increased exercise capacity compared with vehicle-treated controls (**Fig. 3e-i**). These effects were observed without changes in body weight or systolic function, consistent with selective modulation of HFpEF-related phenotypes (**Extended Data Fig. 8**). Together, these findings establish that downstream HGF signaling, rather than HGFAC itself, directly regulates cardiomyocyte hypertrophic remodeling via c-MET activation.

**Fig. 3.**
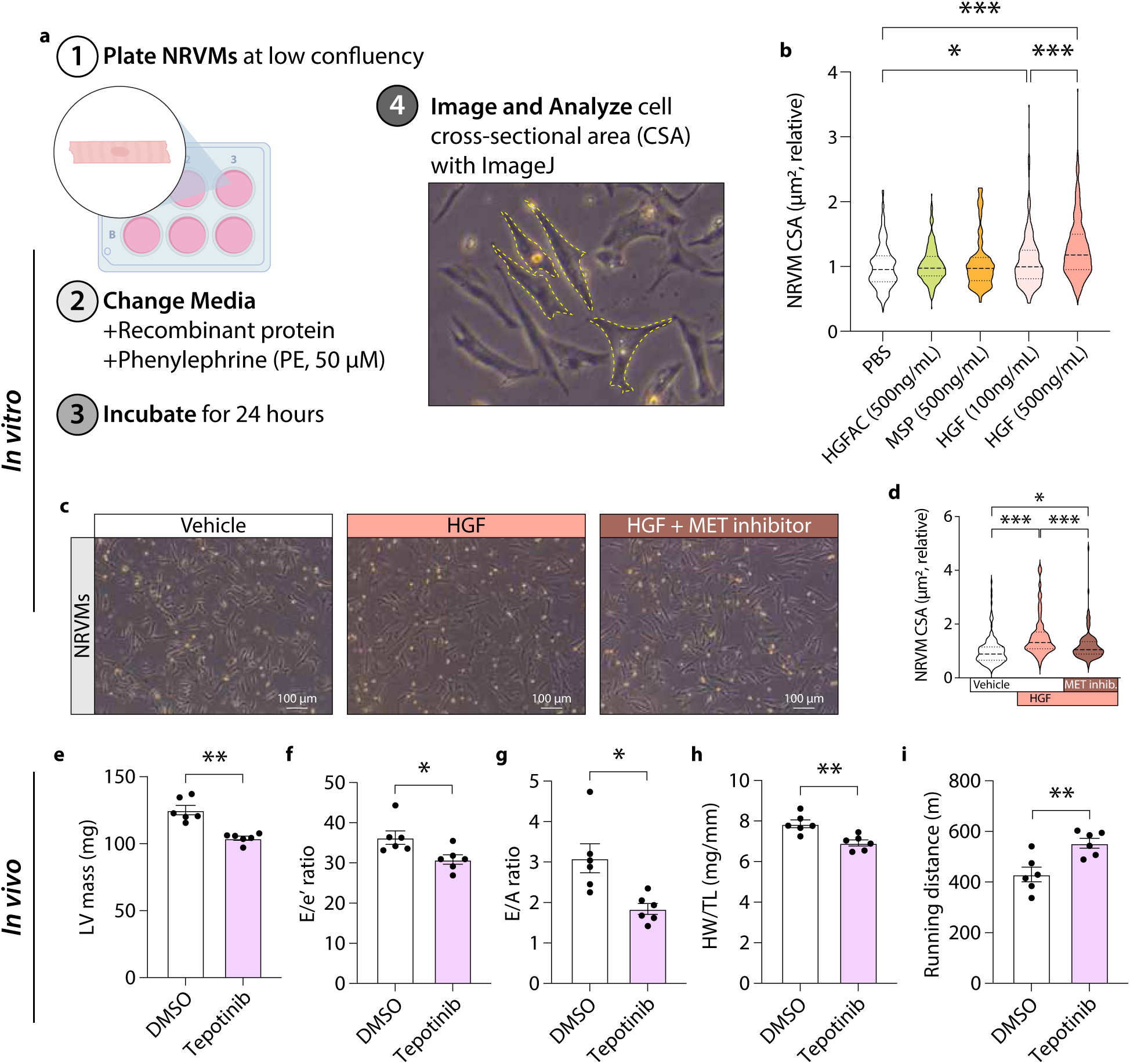
HGF, but not HGFAC, directly induces cardiomyocyte hypertrophy via c-MET signaling, which is blocked by Tepotinib. (**a**) General workflow for cell culture experiments using NRVMs. (**b**) Relative myocyte cell surface areas (CSA) in response to treatment with the indicated recombinant proteins and concentrations. (**c**) NRVMs were treated as in (**a**) with the addition of a Tepotinib at the time of media change. Representative images (left) and quantifications (right) of relative myocyte CSA from the (**c**) Tepotinib study. Each data point represents a cell. n = 3 independent NRVM culture experiments, each performed in technical triplicate with approximately 30 cells quantified per replicate then graphed as violin plots. Data are shown as mean ± SEM. *P < 0.05, **P < 0.01, and ***P < 0.001 by one-way ANOVA. (**e-i**) 8-week-old C57BL/6J male mice were injected with either DMSO or Tepotinib (every other day) and were supplemented HFD and L-NAME for a total of 7 weeks. Assessment of cardiac function using (**e-g**) echocardiography to measure (**e**) LV mass, (**f**) E/e’ ratio, and (**g**) E/A ratio, (**h**) HW/TL, and (**h**) running distance after 7 weeks of HFpEF diet. Each data point represents a mouse (n = 6 mice/group). *P < 0.05 and **P < 0.01 by Student’s t test.

We next assessed whether circulating components of the HGFAC–HGF axis are associated with heart failure in humans. In the UK Biobank^26,27^ proteomic cohort, higher circulating HGF levels were independently associated with increased risk of incident heart failure after adjustment for established cardiovascular risk factors (**Fig. 4b**). This association was validated in the Cleveland Clinic GeneBank^28–33^ cohort (**Supplemental Table 1**) in which elevated HGF levels were associated with HFpEF (**Fig. 4d**). In addition, higher circulating HGF levels were associated with increased odds of elevated cardiac biomarkers, including NT-proBNP and hs-TnT, further supporting a link between HGF signaling and myocardial stress and injury (**Extended Data Fig. 9).** In contrast, circulating HGFAC showed weaker and non-significant associations with heart failure outcomes (**Fig. 4a,c**). Together, these findings suggest that circulating HGF is associated with heart failure risk, including HFpEF, supporting translational relevance of this liver-heart signaling pathway in humans.

**Fig. 4.**
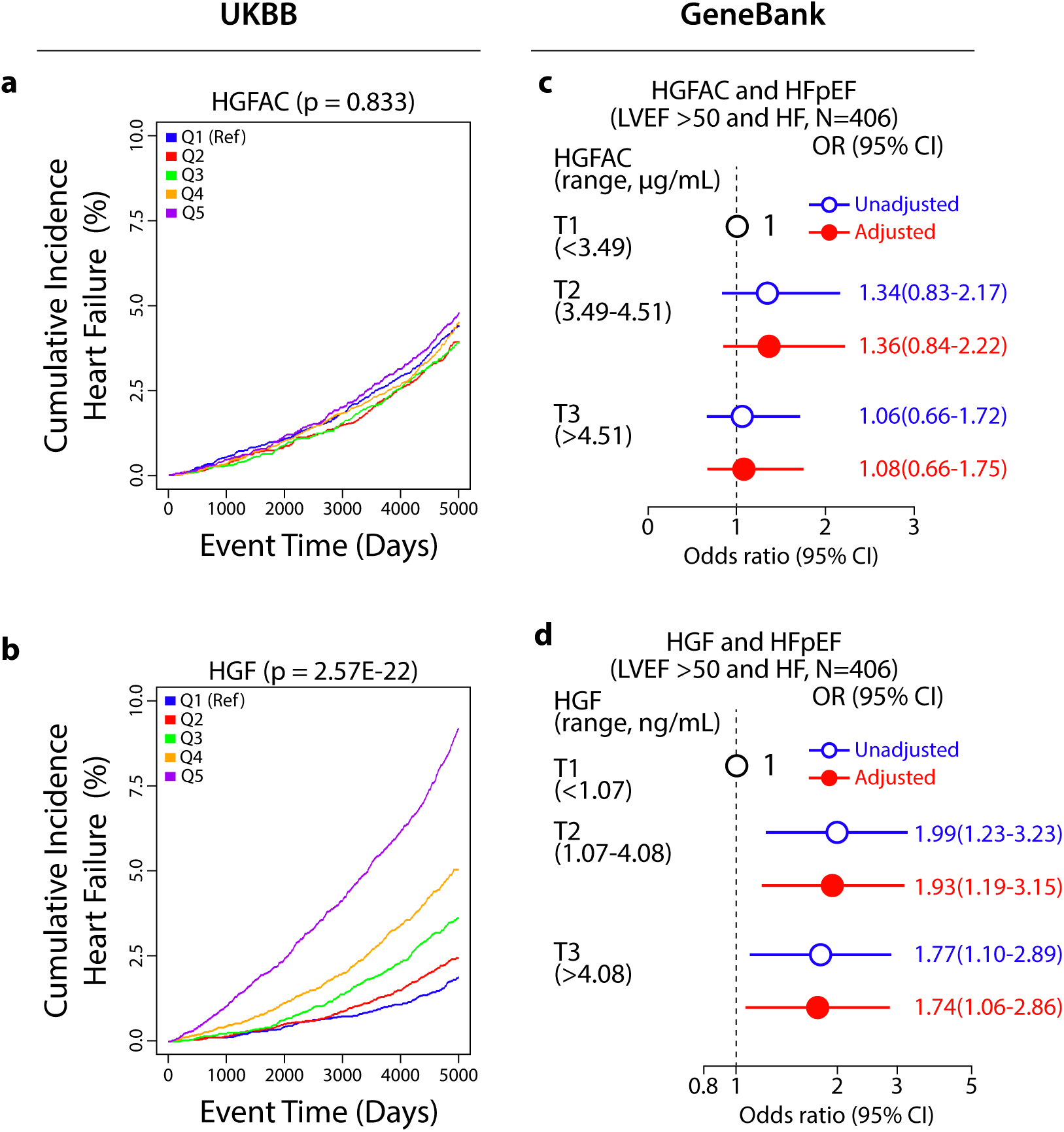
Elevated circulating HGF is associated with increased risk of heart failure in humans. Levels of (**a**) HGFAC and (**b**) HGF were measured in the UK Biobank cohort at baseline (n = 37,055) on the Olink platform. Cox proportional hazards regression models were used to evaluate quintiles of serum HGFAC and HGF protein levels with cumulative incidence of HF with adjustment for age, sex, self-reported ethnicity, BMI, LDL, HDL, systolic blood pressure, diabetes, lipid-lowering medications, and anti-hypertension medications. Forest plots indicating the odds of HFpEF in Cleveland cohort (n = 406) by (**c**) HGFAC and (**d**) HGF tertiles from plasma samples assayed by ELISA. Multivariable logistic regression model for odds ratio included adjustments for age, sex, current smoking, SBP, DM, dyslipidemia (high-density lipoprotein cholesterol ≤40 mg/dL, or low-density lipoprotein cholesterol≥130 mg/dL or triglycerides≥150 mg/dL), symbols represent odds ratios, and the 95% confidence interval is indicated by line length.

In this study, we identify a conserved liver–heart endocrine axis centered on HGFAC and its downstream effector HGF, linking hepatic protease activity to cardiomyocyte remodeling and heart failure. Across complementary systems-level analyses, hepatic HGFAC was associated with coordinated cardiac gene expression programs in both mice and humans, suggesting conserved inter-organ transcriptional coupling. *In vivo*, hepatic HGFAC overexpression exacerbated HFpEF phenotypes, while pharmacologic inhibition of c-MET attenuated disease features, supporting a functional role for downstream HGF signaling in cardiac remodeling. Cardiomyocytes emerged as the principal responding cardiac cell type, consistent with activation of receptor tyrosine kinase signaling pathways. Single-cell analyses further suggested that cardiomyocyte activation is accompanied by broader remodeling of intercellular communication within the ventricle, indicating that perturbation of this axis may contribute to myocardial network organization beyond cell-autonomous effects. The discrepancy between HGFAC and HGF associations in human cohorts likely reflects differences in tissue specificity, post-translational activation, and systemic integration of signaling activity. Whereas HGFAC is predominantly liver-enriched and transcriptionally constrained, circulating HGF may better reflect integrated pathway activity across tissues, which may explain its stronger association with incident heart failure. Together, these findings define a conserved endocrine signaling pathway linking hepatic proteolytic activity to cardiomyocyte remodeling and heart failure susceptibility. More broadly, they highlight inter-organ protease–growth factor signaling as a mechanism through which liver-derived factors influence cardiac disease risk.

Several limitations should be considered. Although c-MET inhibition attenuated HFpEF phenotypes in mice, the long-term safety and tissue specificity of this pathway require further investigation. In addition, human proteomic associations are observational and do not establish causality. Future studies will be needed to determine whether modulation of this axis alters heart failure risk in humans.

## Funding

This work was supported by the National Heart, Lung, and Blood Institute at the National Institutes of Health (grant numbers U54HL170326, DK117850, and R01HL152176 to A.J.L.). H.A. was supported by the National Heart, Lung, and Blood Institute at the National Institutes of Health (grant numbers U54HL170326 and R01HL168493 to H.A.). S.L.H was supported in part by grant P01 HL147823 from the National Institutes of Health (NIH). M.J. was supported by the Ruth L. Kirschstein National Research Service Award at the National Institutes of Health (grant number T32HL069766 to M.J.) and the Center for Duchenne Muscular Dystrophy at UCLA Azrieli Graduate Student Award. D.C.S. was supported by the National Heart, Lung, and Blood Institute at the National Institutes of Health (grant number T32 HL144449 to D.C.S).

## Acknowledgements

We thank In Sook Ahn, Guanglin Zhang, and Graciel Diamante for their help with the single-nuclei study. We also gratefully acknowledge the UK Biobank Resource for providing access to their data under Application Number 33307.

## Methods

### Study approval

All animal experiments were approved by the Institutional Animal Care and Use Committees of the University of California Los Angeles (UCLA) and conducted in accordance with the Guide for the Care and Use of Laboratory Animals. All participants in the UK Biobank provided informed consent, and the study protocol was approved by the North West Multi-Centre Research Ethics Committee and carried out in accordance with the Declaration of Helsinki; the present analyses were approved by the Institutional Review Board of the USC Keck School of Medicine. Written informed consent was obtained from all participants in the GeneBank study, which was approved by the Institutional Review Board of the Cleveland Clinic, and the present analyses were approved by the Institutional Review Board of the David Geffen School of Medicine at UCLA.

### Animals

Mice were housed under a 12-hour light/dark cycle (lights on from 6 a.m. to 6 p.m.) at ambient temperature, with ad libitum access to food and water. Wild-type C57BL/6J mice (Strain No. 000664) were sourced from the Jackson Laboratory. Animals were randomly assigned to treatment groups, and all experiments were conducted using standard randomization and blinding procedures. Sample size was determined empirically based on prior similar studies, with no samples excluded intentionally from analyses. Euthanasia was performed by isoflurane overdose followed by cervical dislocation. After euthanasia, tissues were weighed and frozen in liquid nitrogen.

### Hybrid Mouse Diversity Panel

The 100 strains of inbred mice in the Chow-HMDP, selected based on genetic diversity and other criteria^12,34^, were obtained from the Jackson Laboratory. The mice were fed a chow diet until sacrifice at 16 weeks of age. Following a 16h fast, mice were bled retro-orbitally under isoflurane anesthesia. Mice were euthanized using isoflurane overdose followed by cervical dislocation. Upon euthanasia, tissues were weighed and then frozen in liquid nitrogen. Some tissues, including liver and heart, were subjected to RNA sequencing to determine gene expression levels. Endocrine interactions between the liver and heart were identified using cross-tissue correlation analysis^12,34^. The framework was applied to the following GEO accessions: chow liver – GSE16780 and chow heart – GSE77263.

### Sequence alignment

Sequence alignments made using NCBI COBALT multiple sequence alignment tool (https://www.ncbi.nlm.nih.gov/tools/cobalt/cobalt.cgi?CMD=Web). Sequence identities calculated via ALIGNMENTVIEWER (https://alignmentviewer.org).

### Pathway enrichment analysis

Pathway analysis was performed using a web-based tool named Enrichr^35^ that visualizes enriched terms and genes across multiple selected gene set libraries. The gene set libraries used in this study were GO Molecular Function and NCI-Nature.

### Tissue crosstalk using GD-CAT

The genetically-derived correlations across tissues (GD-CAT)^20^ resource was utilized to perform crosstalk analysis on data from the human Genotype-Tissue Expression (GTEx)^21^ project.

### Genetic variant identification using PhenoScanner

Genetic variants located within or proximal to the *HGFAC* and *HGF* loci were identified using PhenoScanner V2, a curated database of publicly available genome-wide association study (GWAS) results^36^.

### Mouse model of HFpEF

Mice were fed either chow diet (Teklad, Cat. No. 2916) or HFpEF diet (Research Diets, Cat. No. D12492), which contains 60% fat, with Nω-Nitro-L-arginine methyl ester hydrochloride (l-NAME) (Sigma Cat. No. N5751) added to the drinking water at a concentration of 0.5 g/L (pH 7.4). Tepotinib (MedChemExpress, Cat. No. HY-14721) was dissolved in 10% DMSO (Fisher, Cat. No. 67-68-5) and 90% 20% sulfobutylether-β-cyclodextrin (MedChemExpress, Cat. No. HY-17031). The solution was adjusted to the working concentration with 0.9% sodium chloride (Hospira, Cat. No. 0409-4888-02) and injected (i.p.) at 15 mg/kg body weight, three times per week, from the start of the HFpEF diet and continuing until euthanasia at 7 weeks.

### Adeno-associated viruses (AAV)

AAV8-TBG-*Null* and AAV8-TBG-m-HGFAC (AAV-261274) were obtained from Vector Biolabs. Virus was diluted with 0.9% sodium Chloride Injection, USP (Hospira, NDC 0409-4888-02) and then 100 µL of virus was i.p. injected into each mouse at the titer of 5×10^11^ gc/mouse.

### Transaortic echocardiography

Mice were imaged non-invasively. Briefly, the mice were anesthetized and maintained with 2% isoflurane in 95% oxygen. A Vevo 3100 (Visual Sonics) echocardiography system with a 30mHz transducer was used to acquire the images. A parasternal short axis view was recorded. The short axis view was used to generate M-mode images for analysis of ejection fraction and fraction shortening. A modified four-chamber view was recorded and used to generate pulsed wave doppler and tissue doppler images for analysis of mitral valve E/A and E/e’. At the end of the procedure all mice recovered from anesthesia without difficulty.

### Treadmill fatigue test

Mice were trained on the treadmill apparatus for five consecutive days. Following a 24h rest day, testing was performed as described^37^. Trials were terminated when a mouse refused to interact with the treadmill (spending ≥ 10s at the base of the treadmill lane containing the electric-stimulus grid).

### ELISA

HGFAC (BioMatik, Cat. No. EKC37044) and HGF (Bio-techne, Cat. No. DY2207) enzyme-linked immunosorbent assays (ELISA) were performed using mouse plasma samples according to the manufacturer’s recommendations. As above, blood was collected retro-orbitally under isoflurane anesthesia using BD Microtainer (Tubes with K2EDTA, Ref No. 365974) and then plasma was isolated by centrifugation for 10 minutes at 2,000 x g using a refrigerated centrifuge.

### Western blotting

Western Blots were performed using pro-HGFAC (R&D systems, Cat. No. AF1715) antibody. Following transfer, membranes were then blocked in TBST (Santa Cruz Biotechnology, Cat. No. sc-362311) containing 5% skim milk (VWR, Cat. No. 232100). Membranes were placed in primary antibodies on a shaker overnight at 4°C. The following day, membranes were washed in TBST and then placed in secondary antibodies for 1 hour at room temperature. Blots were then washed in TBST and placed in Amersham ECL Prime Western Blotting Detection solution (Sigma, Cat. No. RPN2236). Blots were imaged using an iBright CL750 Imaging System (ThermoFisher, Cat. No. A44116) and bands were quantified using ImageJ Software (National Institutes of Health).

### Single-nucleus RNA sequencing (snRNAseq)

Following isoflurane anesthesia, mouse hearts (n = 1 pool of 3 hearts per group) were excised and the lower ventricular portions were dissected and then flash frozen. All the following steps were performed on ice and under cold conditions unless described otherwise. All solutions described in the following steps contained Protector RNase Inhibitor (Sigma, Cat. No. 3335399001). Frozen ventricle samples (heart) were dissociated using Nuclei EZ lysis buffer (Sigma, Cat. No. NUC101). The homogenate was then spun at 600 x g for 5 minutes at 4°C. The supernate solution containing the lysis buffer was then removed and nuclei were resuspended and pooled in PBS (Corning, 21-040-CM). Samples were then resuspended with Debris Removal Reagent (10X Genomics, PN: 2000560) and spun at 700 x g for 10 minutes at 4°C. The supernate solution was then aspirated, and nuclei were resuspended in PBS. Nuclei were then spun at 600 x g for 5 minutes at 4°C and resuspended based on the predetermined concentrations. Single-nuclei libraries were constructed (10x Genomics Chromium GEM-X Single Cell 3’ Reagent Kits v4, PN-1000686) and sequenced (NovaSeq S4 X Plus 10B). CellRanger (v 8.0) and Seurat (v 5.0) were used for reads alignment, quality control, and cell type-specific differential gene expression analysis. We obtained a combined total of 88,470 nuclei after excluding the droplets containing either doublets or ambient RNA. Annotation of the different cell types was done based on the expression of known marker genes^38^. Non-parametric Wilcoxon rank sum test was used to compare gene expression between treated and control groups. Multiple testing was corrected using the Benjamini-Hochberg method. Quantitative assessment of global transcriptome shifts was done as described elsewhere^39^. To ensure robust comparisons, we included cell types containing a minimum of 1,000 total cells across conditions. The average gene expression of each gene within one cell type was calculated for *Hgfac* and *Null* groups. Euclidian distance in gene expression was calculated as a metric to quantify the effect of the treatment on each cell type. To determine if the observed Euclidean distance between *Hgfac* vs *Null* cells within each cell type was significantly larger than that of random cells, we estimated a null distribution by calculating the Euclidean distance between randomly sampled cells of the given cell type. This permutation approach was repeated 1000 times to generate the null distribution, which was compared to the Euclidean distance generated from the true *Hgfac* and *Null* groups to determine an empirical p-value. To correct for multiple testing across all the cell types tested, we applied a Bonferroni correction to retrieve adjusted p-values.

### Cell culture

Neonatal mouse ventricular myocytes were isolated as reported previously^40^. Cells were isolated from P2–3 pups using serial trypsin/DNase digestions followed by Percoll density gradient centrifugation, and the purified myocytes were collected and resuspended in DMEM/F12 (ThermoFisher, Cat. No. 11330032) plating medium under NIH– and UCLA-approved animal care protocols. NRVMs were resuspended in day 0 plating conditions supplemented with 10% FCS and penicillin/streptomycin. Myocytes were plated on fibronectin (ThermoFisher, Cat. No. 33016015) coated plates. After 24 hours the media were changed to remove FCS which was then replaced with Insulin-Transferrin-Selenium-Ethanolamine (ITS) (ThermoFisher, Cat. No. 51500056) solution. After 48h hours, cells were ready for treatment with recombinant proteins in the absence of FCS and ITS.

Mouse HGFAC (Biotechne, Cat. No. 1200-SE), mouse MSP/MST1 (MedChemExpress, Cat. No. HY-P701103) and mouse HGF (Abcam, Cat. No. ab281797) were used in the NRVM cell culture experiments. Recombinant proteins were added into DMEM/F12 buffer containing penicillin/streptomycin in the presence of phenylephrine hydrochloride (Sigma, Cat. No. P6126). After removal of the supernate containing ITS, NRVMS were then treated with recombinant proteins (500ng/mL unless indicated otherwise) and phenylephrine (50uM) containing media for 24 hours. The cell culture plates were then imaged using brightfield microscopy and images were analyzed for changes in hypertrophy using ImageJ.

The inhibitor c-MET inhibitor Tepotinib (MedChemExpress, HY-14721 was used in the NRVM cell culture experiments. Tepotinib (100nM) was added in the presence of recombinant mouse HGF (500ng/mL) and phenylephrine containing media for 24 hours. The cell culture plates were then imaged using brightfield microscopy and images were analyzed for changes in hypertrophy using ImageJ.

### Clinical Studies

The UK Biobank is a large, multi-site cohort consisting of recruited participants between 40-69 years of age who were registered with a general practitioner of the UK National Health Service. Between 2006-2010, a total of 503,325 individuals were enrolled through 22 assessment centers in the UK^26^. Extensive data on demographics, ethnicity, education, lifestyle indicators, imaging of the body and brain, and disease-related outcomes were obtained at enrollment through questionnaires, health records, and/or clinical evaluations. Baseline serum samples from a subset of 54,219 participants were also used for proteomics profiling of ∼3000 proteins with the Olink platform^27^.

Time-to-event analyses in the UK Biobank were carried out to test serum HGFAC and HGF protein levels for association with incident heart failure based on participants being assigned International Classification of Diseases version-10 (ICD10) codes I15.0, I15.1, or I15.9 (Data-Field 41270 and 41280) after enrollment into UK Biobank and up to October 31, 2022 (maximum follow-up period was capped at 5,000 days). Only participants who did not have an ICD10 code or self-reported heart failure (Data-Field 20002) in their records at baseline and had full clinical data (n = 37,055) were included in these analyses, of which 1,514 developed heart failure over 5000 days of follow-up. Cox proportional hazards regression models were used to evaluate quintiles of serum HGF and HGFAC protein levels with incident risk of HF with adjustment for age, sex, self-reported ethnicity, BMI, LDL, HDL, systolic blood pressure, diabetes, lipid-lowering medications, and anti-hypertension medications. Results of time-to-event analyses are reported as hazard ratios (HR) with 95% confidence intervals (CI) and quintile comparison P-values. All analyses were carried out with in R (v4.3.0, R Core Team, Vienna, Austria).

The Cleveland Clinic GeneBank study is a single-site sample repository comprised of ∼10,000 sequential consenting stable subjects undergoing elective diagnostic cardiac evaluations with extensive clinical and laboratory characterization and adjudicated longitudinal observation (<u>clinicaltrials.gov</u> identifier: NCT00590200). Participant recruitment occurred between 2001 and 2007 from a large geographic catchment area with significant out of state enrollment (48 of 50 US states represented amongst enrolling subjects). Exclusion criteria for GeneBank included myocardial infarction (MI) within the preceding four weeks of enrollment, or elevated troponin I (>0.03 mg/dL; Abbott Architect) at enrollment. Ethnicity was self-reported, and information regarding demographics, medical history, and medication use was obtained by patient interviews and confirmed by chart reviews at baseline enrollment. All clinical outcome data were verified by source documentation. The GeneBank Study has been used previously for discovery and replication of novel genes and risk factors for cardiovascular outcomes^28–33^.

Studies were performed on a subset of 203 GeneBank subjects with HFpEF, defined as having a clinical diagnosis of HF but with left ventricular ejection fraction (LVEF) ≥ 50%, and 203 control subjects with LVEF ≥ 50%, and without HF (NHF). Baseline characteristics of the subjects in the Cleveland Clinic cohort were listed in **Supplementary Table 1**. Human HGFAC (Bio-techne, Cat. No. DY1514) and HGF (Bio-techne, Cat. No. DY294) solid phase sandwich ELISAs were performed according to the manufacturer’s recommendations. The samples were run in duplicate with standards and 2 quality control samples on each plate to account for plate-to-plate variation.

The plates were analyzed with Molecular Devices SpectraMax 190 and analyzed with Softmax PRO software for quantification relative to the plate-specific standard curve. The coefficients of variation (CVs) for two quality-control (QC) samples were 10.8% and 15.8% for HGFAC, and 6.2% and 9.8% for HGF. Odds ratio (OR) and corresponding 95% confidence intervals (CI) for HFpEF were calculated with both univariable (unadjusted) and multivariable (adjusted) logistic regression models according to HGF tertiles and HGFAC tertiles. The logistic regression model was adjusted for traditional cardiac risk factors including age, sex, current smoking, systolic blood pressure, diabetes mellitus, and dyslipidemia (defined as HDL≤40 mg/dL, LDL≥130 mg/dL, or triglycerides ≥150 mg/dL). All analyses were performed using R, version 4.3.3 (R Project for Statistical Computing), and 2-sided P values <0.05 were considered statistically significant.

### Statistics

All computational procedures were performed using R statistical software. Correlations and associated p-values were calculated using biweight midcorrelation. Single comparisons between two groups were conducted using two-tailed Student’s t-tests with 95% confidence intervals. Multiple comparisons were analyzed using a one-way ANOVA followed by Tukey’s multiple comparisons test, or a two-way ANOVA followed by Sidak’s multiple comparisons test. A significance level of p < 0.05 was considered statistically significant. Unless otherwise noted, values are expressed as means ± SEM. Biweight midcorrelation coefficients and p-values for liver-heart cross-tissue analysis were calculated using the R package Weighted Gene Co-expression Network Analysis (WGCNA). Genome-wide association analysis of clinical traits and liver/heart gene expression data was conducted using FaST-LMM, with a significance threshold of 3.46 x 10^-6^ determined by permutation and modeling. Linkage disequilibrium (LD) was assessed by calculating pairwise r² SNP correlations for each chromosome

## Data availability

All data that support the conclusions in this manuscript can be found in the main text or supplementary materials. The HMDP framework and sequencing data were applied to or deposited in the following GEO accessions: chow liver – GSE16780, chow heart – GSE77263, and single-nuclei RNA sequencing data from Fig. 2 – GSE318505.

## Figures

**Extended Data Fig. 1.**
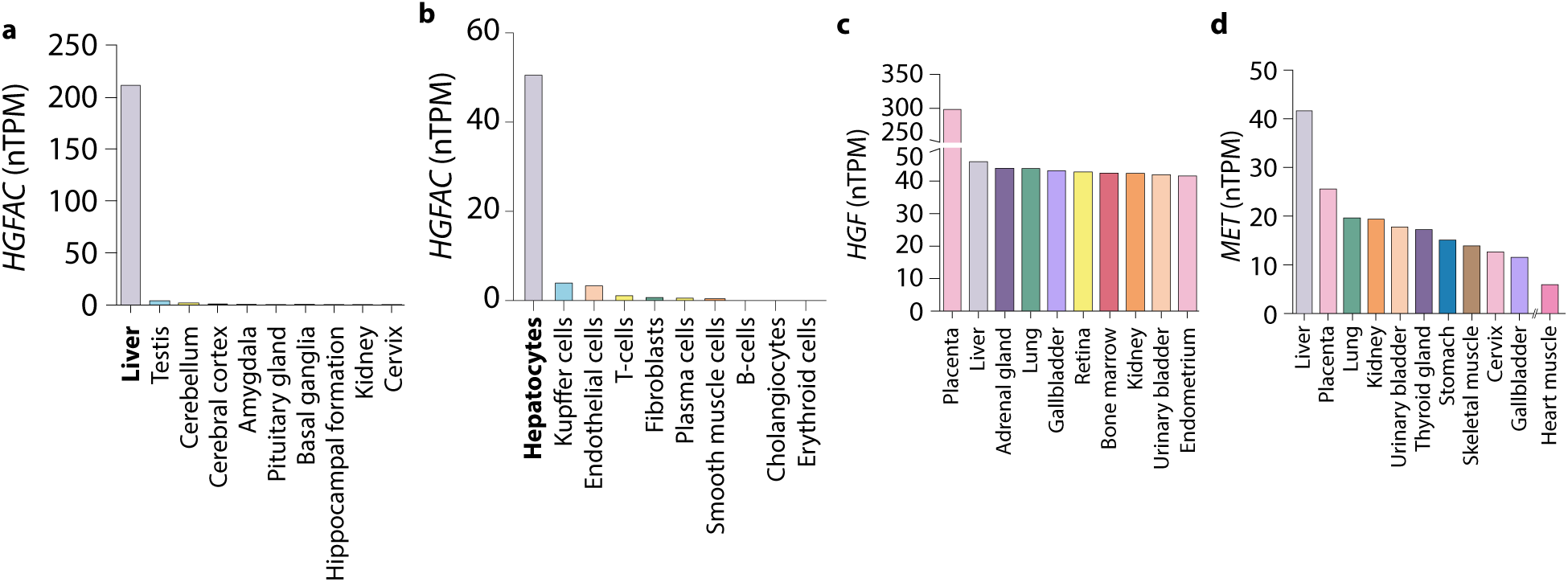
Tissue-specific expression of HGFAC, HGF, and MET in humans. Expression across tissues from The Human Protein Atlas for (**a and b**) *HGFAC*, (**c**) *HGF*, and (**d**) *MET* reported as normalized protein-coding transcripts per million (nTPM).

**Extended Data Fig. 2.**
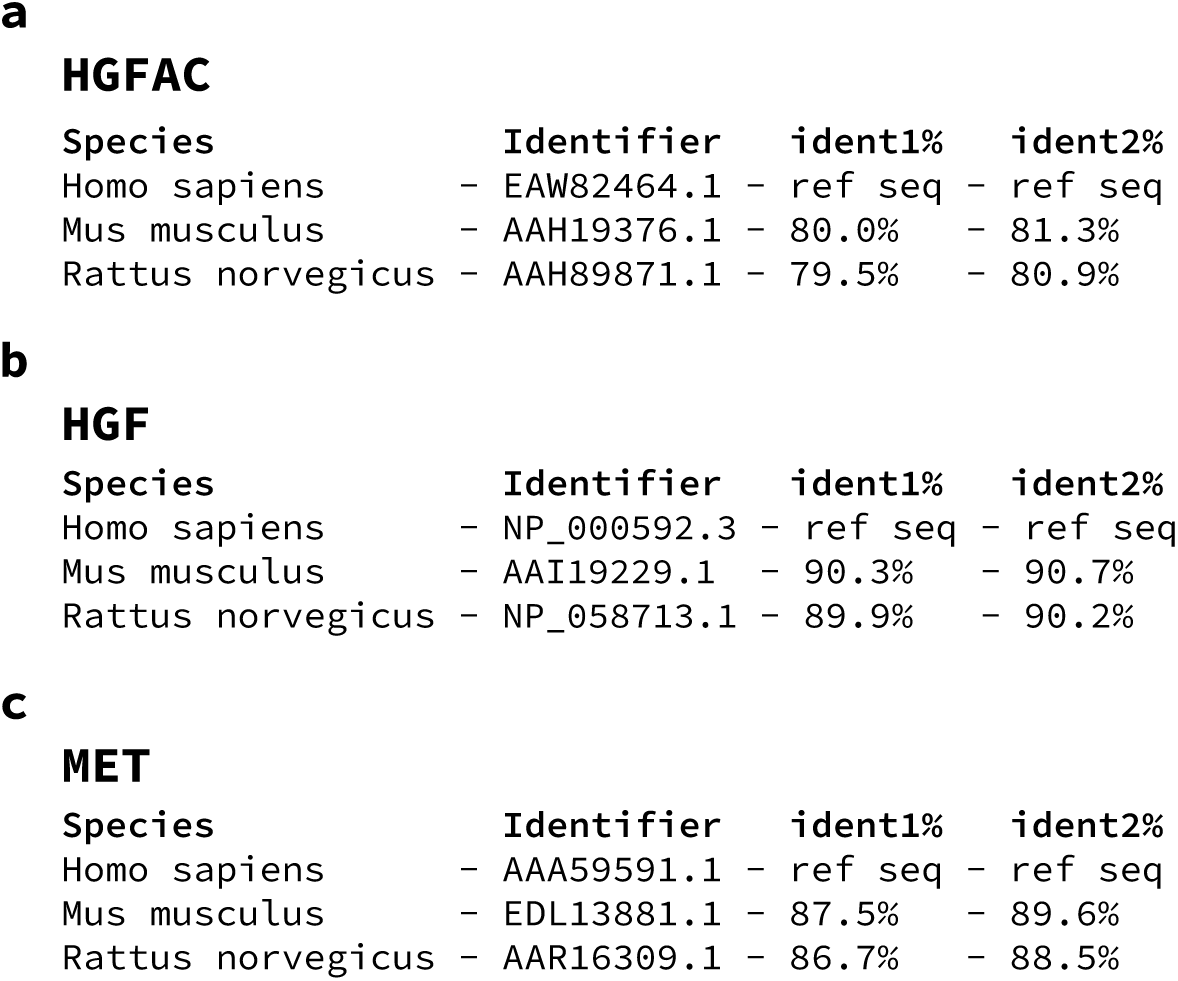
HGFAC, HGF, and MET are conserved across mouse, rat, and human. Protein sequence alignment of (**a**) HGFAC, (**b**) HGF, and (**c**) MET. Species, protein identifier, and percent identities for sequence alignments. ident1% = fraction of reference sequence identical to second sequence (gaps not counted). ident2% = fraction of second sequence identical to reference sequence (gaps not counted).

**Extended Data Fig. 3.**
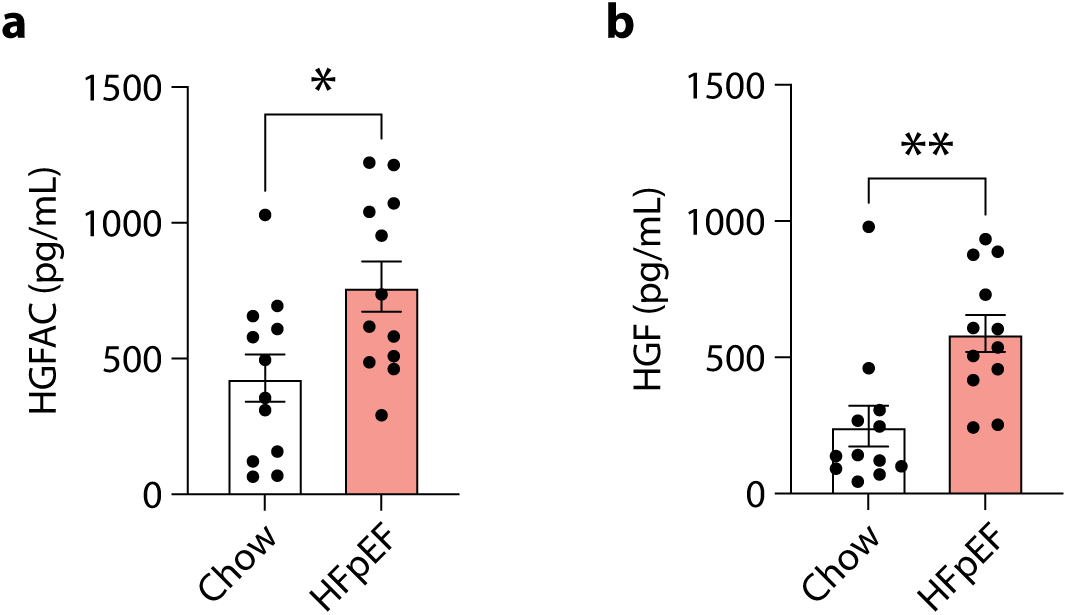
Increased circulating HGFAC and HGF in a HFpEF Mouse Model. ELISA analysis of (**a**) HGFAC and (**b**) HGF in mouse plasma. Data are shown as mean ± SEM. *P < 0.05 and **P < 0.01 by Student’s t test.

**Extended Data Fig. 4.**
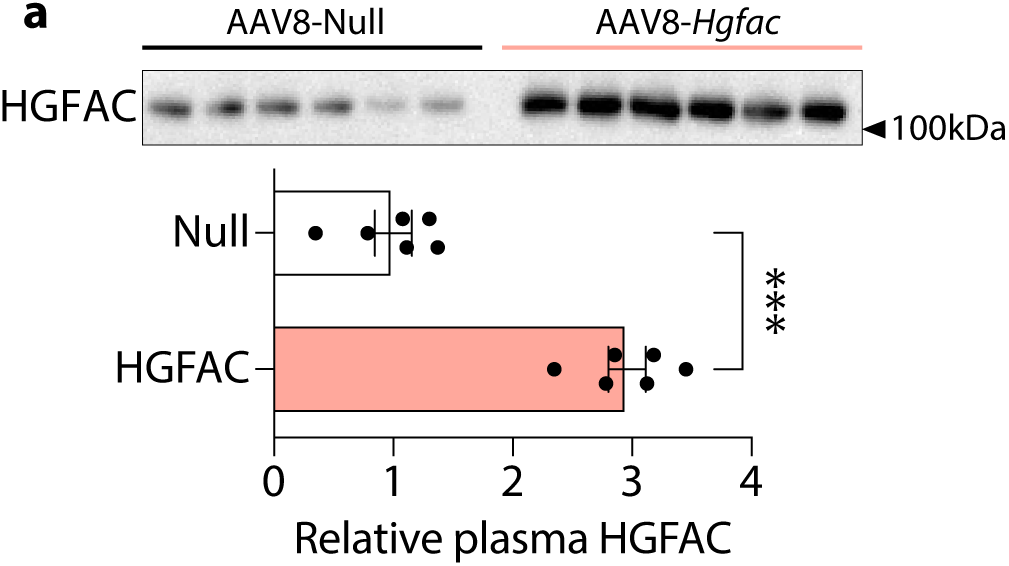
Hepatic *Hgfac* overexpression elevates circulating HGFAC levels. 8-week-old C57BL/6J male mice were injected with adeno-associated virus (AAV8-*Null* or AAV8-TBG-*Hgfac*). (**a**) Western blot imaging (top) and quantification (bottom) show relative HGFAC plasma protein levels.

**Extended Data Fig. 5.**
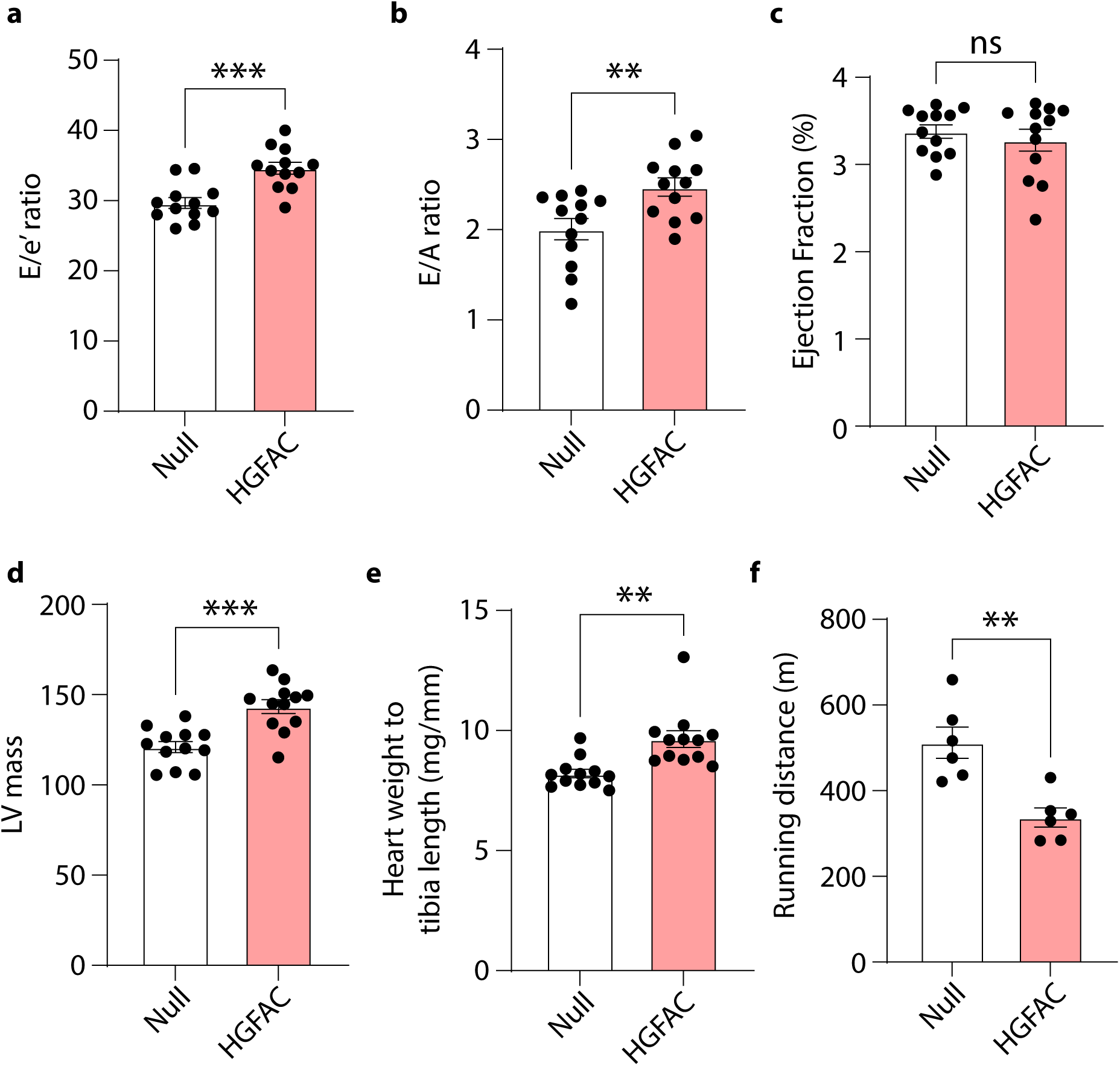
Hepatic *Hgfac* overexpression exacerbates HFpEF-associated cardiac dysfunction in mice. 8-week-old C57BL/6J male mice were injected with adeno-associated virus (AAV8-*Null* or AAV8-TBG-*Hgfac*). Measurements of (**a**) E/e’ ratio, (**b**) E/A ratio, (**c**) Ejection Fraction, (**d**) LV mass, (**e**) Heart weight to tibia length, and (**f**) Running distance after 7 weeks of overexpression and HFpEF diet. All data are presented as mean ± SEM. ns, not significant. *P < 0.05, **P < 0.01, and ***P < 0.001 by Student’s t test.

**Extended Data Fig. 6.**
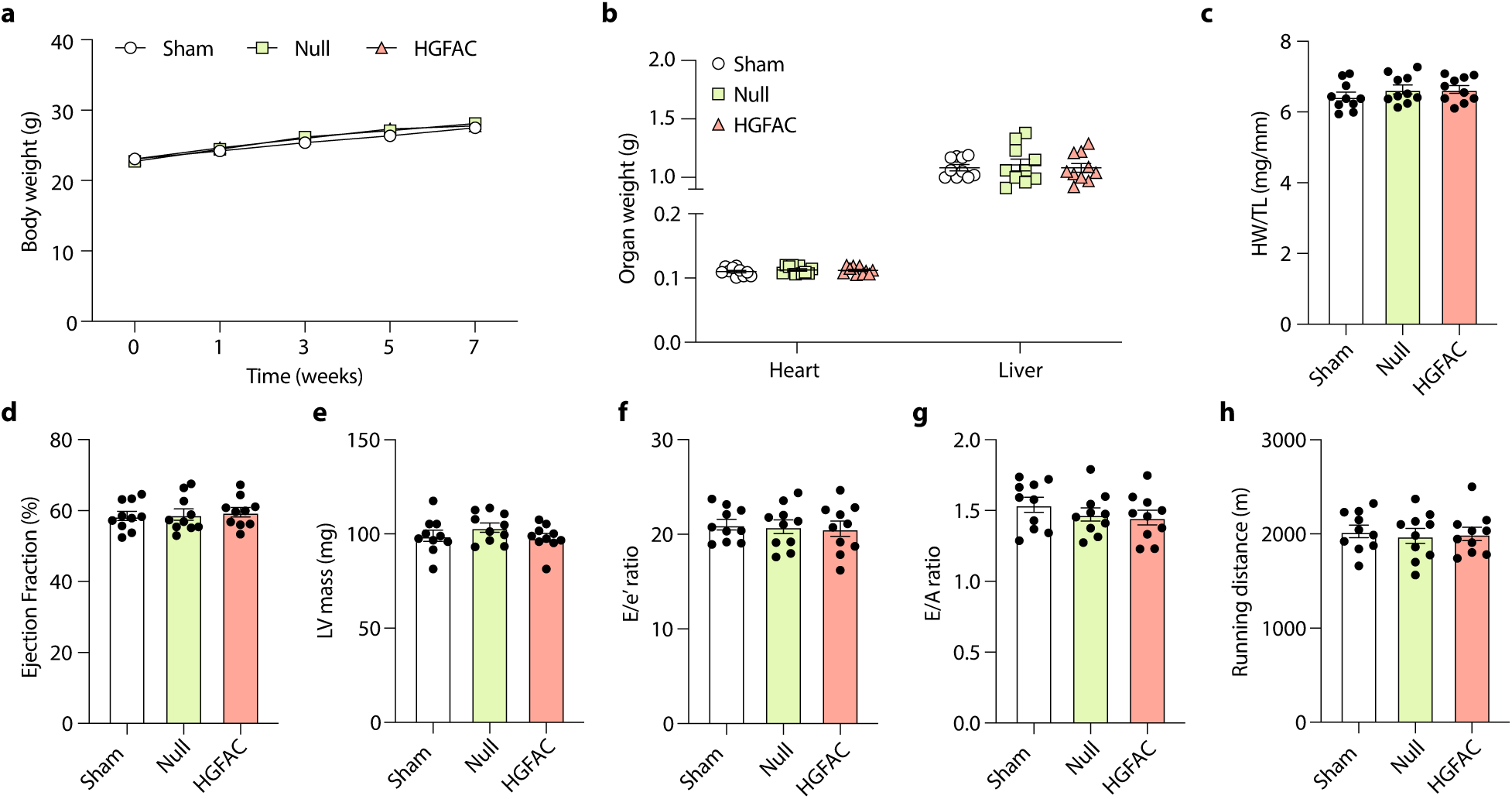
Hepatic *Hgfac* overexpression failed to induce HFpEF-associated cardiac dysfunction in mice on a chow diet. 8-week-old C57BL/6J male mice (n = 10 / group) were injected with saline, AAV8-*Null* or AAV8-TBG-*Hgfac* and were assessed for (**a**) body weight, (**b**) organ weight, (**c**) HW/TL, (**d**) Ejection fraction, (**e**) LV mass, (**f**) E/e’ ratio, (**g**) E/A ratio, and (**h**) running distance. All data are presented as mean ± SEM. ns, not significant.

**Extended Data Fig. 7.**
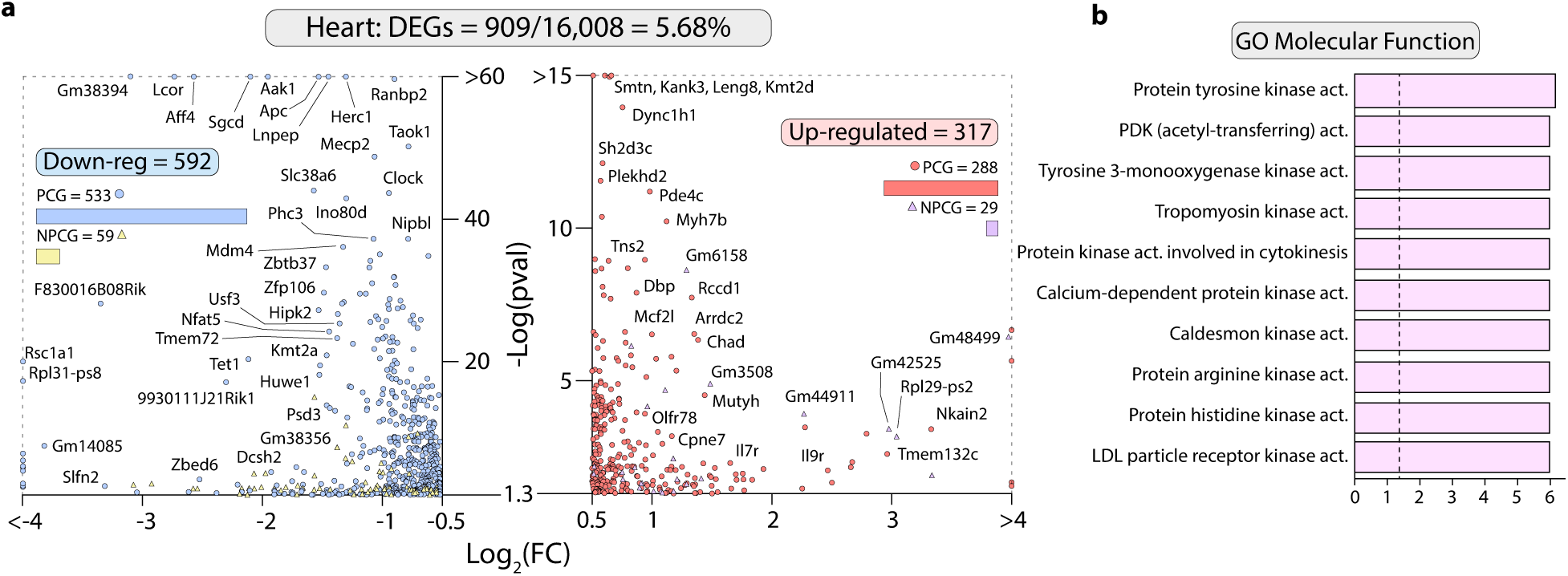
8-week-old C57BL/6J male mice were injected with of AAV8-TBG-*Hgfac* or AAV8-*Null* and were supplemented HFD and L-NAME for 7 weeks. (**a**) Bulk RNA-seq results of significantly (p < 0.05 and Log2FC > 0.5) downregulated (left) and upregulated (right) genes from heart tissue. (**b**) Enrichr-derived ontology analysis using the top 500 most significant DEGs from (**a**).

**Extended Data Fig. 8.**
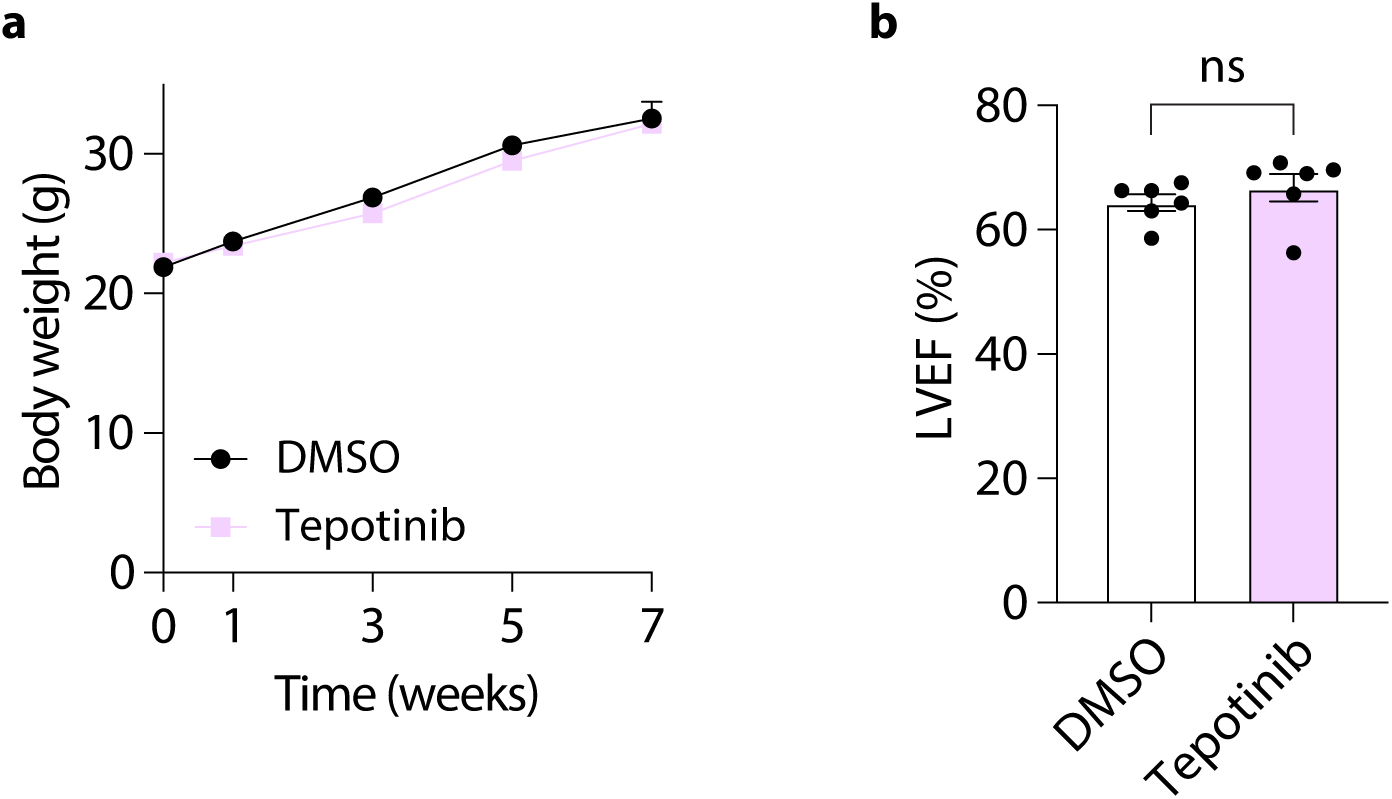
c-MET inhibition with Tepotinib preserves ejection fraction without affecting body weight in HFpEF mice. Assessment of (**a**) body weight and (**b**) cardiac function using echocardiography to measure Ejection Fraction. Each data point represents a mouse (n = 6 mice/group). ns, not significant by Student’s t test.

**Extended Data Fig. 9.**
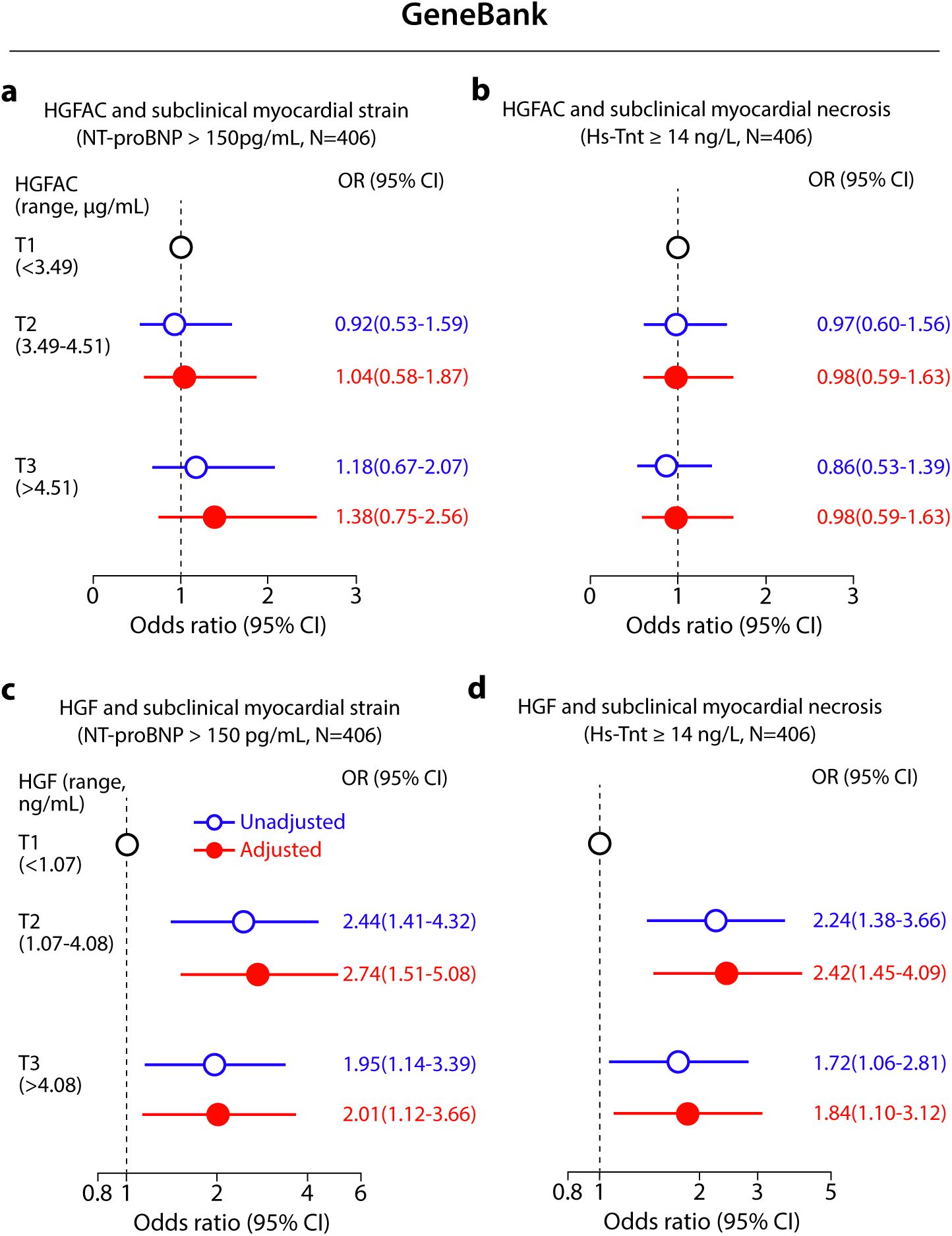
The relationship of HGFAC and HGF with subclinical myocardial strain and necrosis in the Cleveland cohort. (**a and c**) Forest plots depicting the odds of subclinical myocardial strain (NT-proBNP>150 pg/ml) by (**a**) HGFAC and (**c**) HGF tertiles; (**b and d**) Forest plots indicating the odds of subclinical myocardial necrosis (Hs-TnT≥14 ng/L) by (**b**) HGFAC tertiles and (**d**) HGF tertiles. Multivariable logistic regression model for odds ratio included adjustments for age, sex, current smoking, SBP, DM, dyslipidemia (high-density lipoprotein cholesterol ≤40 mg/dL, or low-density lipoprotein cholesterol≥130 mg/dL or triglycerides≥150 mg/dL), symbols represent odds ratios, and the 95% confidence interval is indicated by line length.

**Supplemental Table 1.** Baseline characteristics of subjects in Cleveland cohort.

| Characteristics | All Subjects (N=406) | Subjects without HFpEF (N=203) | Subjects with HFpEF (N=203) | P |
| --- | --- | --- | --- | --- |
| Age, years | 68.9 (60.8- 74.8) | 68.7 (60.7- 74.4) | 69.0 (60.8- 74.9) | 0.82 |
| Male sex (%) | 44.8 | 44.3 | 45.3 | 0.92 |
| Diabetes mellitus (%) | 38.2 | 39.4 | 36.9 | 0.68 |
| BMI (kg/m<sup>2</sup>) | 29.3 (25.0- 33.9) | 29.1 (25.1- 33.2) | 29.4 (25.0- 34.9) | 0.45 |
| Systolic blood pressure (mmHg) | 135(121- 149) | 136(122- 148) | 130 (121- 150) | 0.45 |
| Hypertension (%) | 72.1 | 65.7 | 78.5 | <0.01 |
| Current smoking (%) | 8.4 | 7.4 | 9.4 | 0.58 |
| CAD (%) | 63.1 | 62.6 | 63.5 | 0.92 |
| CVD (%) | 64.5 | 63.5 | 65.5 | 0.76 |
| History of MI (%) | 30.0 | 26.0 | 34.0 | 0.11 |
| LDL cholesterol (mg/dL) | 95.0 (79.0- 114.0) | 95.5 (81.0- 113.0) | 94.0 (76.8- 114.5) | 0.22 |
| HDL cholesterol (mg/dL) | 35.3 (28.4- 42.8) | 35.5 (29.5- 42.4) | 34.9 (27.1- 43.3) | 0.19 |
| Total cholesterol (mg/dL) | 161.0 (139.9- 187.9) | 161.3 (142.0- 186.6) | 159.2 (137.2- 189.4) | 0.39 |
| Triglycerides (mg/dL) | 118.0 (86.0- 172.0) | 117.5 (86.3- 170.8) | 118.5 (86.0- 177.0) | 0.90 |
| CRP (mg/L) | 3.38 (1.37- 7.49) | 2.88 (1.24- 7.13) | 3.66 (1.78- 8.09) | 0.04 |
| eGFR (mL/min/1.73m<sup>2</sup>) | 83.9 (66.8- 94.3) | 88.3 (72.8- 97.0) | 78.6 (61.5- 92.0) | <0.001 |
| LVEF (%) | 55 (55- 60) | 60(55- 65) | 55(55- 60) | <0.01 |
| NT-proBNP (pg/ml) | 345.2 (151.3- 943.9) | 227.6 (90.7- 508.4) | 702.9 (263.3- 1624.5) | <0.001 |
| Hs-TnT (ng/L) | 14.0 (7.3- 22.9) | 10.9 (5.6- 18.8) | 17.0 (9.3- 28.9) | <0.001 |
| Statins (%) | 51.5 | 55.2 | 47.8 | 0.16 |
| Aspirin (%) | 66.3 | 76.4 | 56.2 | <0.001 |
| Anti-diabetic drugs (%) | 21.9 | 19.7 | 24.1 | 0.34 |
| ACE inhibitors (%) | 49.5 | 41.4 | 57.6 | 0.001 |
| Calcium channel blockers (%) | 22.2 | 15.8 | 28.6 | 0.003 |
| Diuretics (%) | 37.7 | 16.3 | 59.1 | <0.001 |
| HGF (ng/mL) | 1.96(0.74- 5.65) | 1.46(0.60- 5.26) | 2.50 (1.00- 6.64) | <0.01 |
| HGFAC (µg/mL) | 3.93 (3.32- 4.73) | 3.94(3.31- 4.73) | 3.92(3.34- 4.72) | >0.99 |

Continuous data are presented median (interquartile range), categorical variables are presented as %. Wilcoxon-rank sum test for continuous variables and $\chi^2$ test for categorical variables were used to examine the difference between groups.

## Notes

### Author Declarations

All participants in the UK Biobank provided informed consent, and the study protocol was approved by the North West Multi-Centre Research Ethics Committee and carried out in accordance with the Declaration of Helsinki; the present analyses were approved by the Institutional Review Board of the USC Keck School of Medicine. Written informed consent was obtained from all participants in the GeneBank study, which was approved by the Institutional Review Board of the Cleveland Clinic, and the present analyses were approved by the Institutional Review Board of the David Geffen School of Medicine at UCLA.

